# How is the distribution of psychological distress changing over time? Who is driving these changes? Analysis of the 1958 and 1970 British birth cohorts

**DOI:** 10.1101/2021.05.31.21257562

**Authors:** Dawid Gondek, Rebecca E. Lacey, Dawid G. Blanchflower, Praveetha Patalay

## Abstract

**Aims:** The main objective of this study was to investigate distributional shifts underlying observed age and cohort differences in mean levels of psychological distress in the 1958 and 1970 British birth cohorts.

**Methods:** This study used data from the 1958 National Child Development Study and 1970 British birth cohort (n=24,707). Psychological distress was measured by the Malaise Inventory at ages 23, 33, 42 and 50 in the 1958 cohort and 26, 34, 42 and 46-48 in the 1970 cohort.

**Results:** The shifts in the distribution across age appear to be mainly due to changing proportion of those with moderate symptoms, except for midlife (age 42-50) when we observed polarisation in distress – increased proportions of people with no or multiple symptoms. The elevated levels of distress in the 1970 cohort, compared with the 1958 cohort, appeared to be due to an increase in the proportion of individuals with both moderate and high symptoms. For instance, at age 33/34 34.2% experienced no symptoms in 1970 compared with 54.0% in the 1958 cohort, whereas 42.3% endorsed at least two symptoms in the 1970 cohort vs 24.7% in 1958.

**Conclusions:** Our study demonstrates the importance of studying not only mean levels of distress over time, but also the underlying shifts in its distribution. Due to the large dispersion of distress scores at any given measurement occasion, understanding the underlying distribution provides a more complete picture of population trends.

## Introduction

Psychological distress captures a continuous distribution of symptoms of depression and anxiety [1]. Examining changes in mental health across age groups and between generations is an important endeavour in research on population health, as it helps to identify vulnerable life stages for mental ill-health and elucidate whether these are changing between generations. Albeit the large heterogeneity of distress across the distribution at the population level [2, 3], most research investigating lifecourse development (age effects) and cross-cohort trends (cohort effects) have focused on mean levels of distress [4-9].

Previous research focussing on trajectories of mean distress suggests that average levels of distress may be somewhat elevated in mid-20s and midlife (age 45-55) [4, 7, 9, 10], and that those born more recently are more likely to experience higher levels of distress throughout adulthood [4, 8, 9]. It is unclear, however, whether the distribution of distress changed across age groups and birth cohorts, and if these changes were driven by specific symptoms. Looking only at the population mean may mask differences in trends within the distribution, and this equally applies to age and cohort effects. For instance, when the proportion of those with high and low distress increases with age or across cohorts, considering means only would suggest a stable trend over time, hiding the more warranted conclusion of increased polarisation in the distribution. On the other hand, an increase in mean distress may be driven by individuals across the entire distribution (i.e., the whole distribution shifts right), or only by those with high levels of symptoms (i.e., change in the shape of the distribution). Identifying subpopulations that drive these changes would help to priorities the allocation of resources to these groups. To our knowledge, only one previous study considered distributional shifts and focused on late adolescence, finding polarisation between those at extreme ends of the spectrum of distress among those aged 16-24 years old in the British Household Panel Survey [11].

Moreover, it is unclear whether higher psychological distress in the 1970 British birth cohort, compared with 1958, is mainly due to poorer mental health in any specific socio-demographic groups. We focused on gender and socioeconomic characteristics, due to being of great policy interest as important determinants of population mental health, with women and those in disadvantaged socioeconomic groups experiencing greater distress [12, 13].

The main objective of this study was to investigate distributional shifts underlying observed age and cohort differences in mean levels of psychological distress across adulthood in the 1958 and 1970 British birth cohorts. This was done both by studying cross-sectional and longitudinal heterogeneity in the distribution. Further, we examined whether observed distributional shifts were driven by specific socio-demographic subgroups, according to gender, parental social class at birth, and highest achieved qualification by age 30/33. Finally, we compared cohort and age differences in the distribution of individual symptoms of psychological distress to examine whether age or cohort trends were driven by any specific symptoms.

## Methods

### Sample

The 1958 birth cohort (National Child Development Study; NCDS) follows the lives of 17,415 people born in England, Scotland, and Wales in a single week of March 1958 [14] and the 1970 British birth cohort (the British birth cohort; BCS70) follows 17,196 people born in England, Scotland, and Wales in a single week of April 1970 [15]. Both cohorts include information on physical and educational development, a range of bio-measures, economic circumstances, employment, family life, health behaviours, wellbeing, social participation, and attitudes [14-16].

Our analytical sample (n=24,707) included those who had at least one measure of distress, were still alive and were not permanent emigrants from Britain by age 50 in the 1958 cohort (n=13,250) and by age 46-48 in the 1970 cohort (n=11,457) (see eFigure 1 for the sample flow diagram). Both 1958 and 1970 cohorts were granted ethical approval for each sweep from 2000 by the National Health Service Research Ethics Committee and all participants have given informed consent.

### Measure of psychological distress

Psychological distress was measured by the Malaise Inventory at ages 23, 33, 42 and 50 in the 1958 cohort and 26, 34, 42 and 46-48 in the 1970 cohort [17]. The 9-item version with “yes-no” response option was used across ages and cohorts (see eTable 1 for items of the questionnaire), with the total distress score ranging from 0-9, where a higher score indicates greater distress.

The Malaise Inventory has good psychometric properties [18], it has been used in the general population and high-risk groups [19]. The measure is suitable for the main objectives of this study, which involves comparing distress scores across age, cohorts, and socio-demographic groups, due to its previously found scalar invariance [20]. Scalar invariance implies that symptoms captured by the items of the Malaise Inventory were interpreted equivalently by participants, regardless of their age (23-50), cohort membership (1958 vs 1970), gender (man vs woman), social class or measurement modes (face-to-face interview vs self-administered questionnaire etc.) [8, 21].

### Demographic variables

Variables used for comparing the demographic composition of subgroups of trajectories of psychological distress were gender, social class at birth, and highest qualification obtained by age 30/33. Gender (man and woman) was based on sex identified at birth by a midwife. Social class at birth represented father’s occupation, coded according to the Register General’s classification, and was categorised as “low” (IV partly skilled/V unskilled), “medium” (III skilled non-manual/manual), or “high” (I professional/II intermediate). The highest qualification obtained by age 30/33 refers to vocational or occupational qualifications (National Vocational Qualifications – NVQ) obtained since school, categorised as “low” (none-NVQ 1), “medium” (NVQ 2-3), or “high” (NVQ 4-5).

### Analytical strategy

First, we described the cross-sectional heterogeneity in the distribution across age and cohorts (1958 vs 1970) using overlying histograms, which allow us to present the extent to which the distributions of distress overlap across cohorts at each age.

Second, we compared the proportion of participants based on their cross-sectional scores of distress, as having 0 symptoms, 1-3 symptoms, and ≥4 symptoms. A score of four or more on the Malaise Inventory is typically considered as indicating a high-risk group for depressive disorder [22-24].

Third, we studied heterogeneity in the longitudinal distribution of psychological distress within each cohort. We used multilevel regression analysis to model cohort-stratified average trajectories of psychological distress across age 23-50 and describe the between and within-subjects variance around these trajectories (see eAppendix 2 for details on the modelling strategy). This method allows for modelling data that are unbalanced in time and include missing data, while accounting for a hierarchical dependency of observations (level 1) within individuals (level 2) – with age becoming an observation-level variable [25].

Fourth, as we identified a large variance around average trajectories, we conducted a latent class growth analysis (LCGA). This approach assumes that observed data consists of a finite number of latent classes, characterised by similar longitudinal trajectories of psychological distress in terms of their intercept and slope [26]. This allowed us to identify subgroups of individuals who shared highly comparable trajectories of distress between age 23 and 50 and compare the distribution and characteristics of these groups across both cohorts. LCGA was conducted using pooled samples from both cohorts. Models with an increasing number of classes, from two to five, were estimated to identify the optimum solution. Alternative specifications were compared based on model fit indicators – Akaike Information Criterion (AIC), Bayesian Information Criterion (BIC), adjusted Bayesian Information Criterion (adj BIC), (Vuong) Lo–Mendell–Rubin likelihood ratio tests ((V)LMR-LRT) – entropy, the class size (>5%) and interpretability of the classes [26, 27].

Subsequently, due to high entropy (>0.80) – participants were allocated to different classes according to their posterior probabilities [27]. Fifth, we used robust Poisson regression to compare the demographic composition of each class across cohorts (1958 vs 1970), gender, social class at birth, and highest qualification obtained by age 30/33 [28]. Differences in the likelihood of being in each class were expressed in relative terms as a risk ratio. Separate regression analyses were run for each demographic exposure, with a dummy indicator of each class being an outcome. Any differences between the cohorts in the demographic composition of classes were tested, with Wald tests, by including an exposure*cohort interaction term in the regressions.

Finally, to examine whether any distributional shifts were driven by specific symptoms captured by the Malaise Inventory, we descriptively compared the proportion of cohort members endorsing individual symptoms of the Malaise Inventory across age and cohorts.

Missing information in multilevel regression analysis and LCGA was accounted for by using Full Information Maximum Likelihood (FIML) [29]. Missing information on individual symptoms and demographic variables used in Poisson regression were multiply imputed, resulting in 20 datasets (see eAppendix 1 for more details). LCGA was conducted using MPlus Version 8 [30], all other analyses were run using Stata 16 [31].

## Results

### Cross-sectional distribution of psychological distress

#### Age differences within each cohort (1958 and 1970)

The mean score of psychological distress was highest at age 42 and 50 (mean at both ages: 1.54) in the 1958 cohort, and at age 42 (mean: 1.90) in the 1970 cohort (see Figure 1). The spread of the distribution increased with age, to a greater extent in the 1970 cohort, being largest at age 50 in the 1958 cohort (standard deviation: 1.98) and at age 46-48 in 1970 (standard deviation: 2.17) (see Figure 1 and eFigure 2).

**Figure 1.**
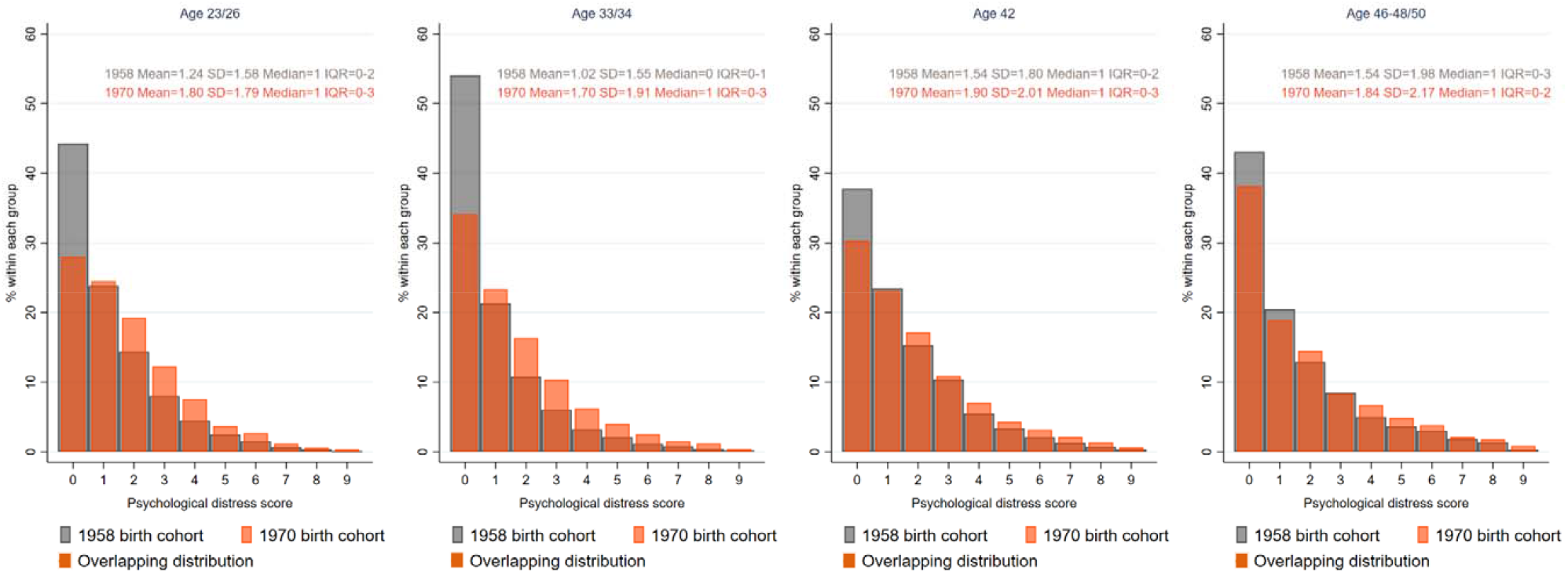
Distribution of psychological distress scores across age and cohorts. *Note*. The figure represents a cross-sectional distribution of psychological distress at each age across cohorts. In addition, mean, standard deviation (SD), median, and interquartile range (IQR) are provided.

As presented in Figure 2, there was a modest decline in the proportion of those with 4+ symptoms (considered as high levels of distress) between age 23/26 and 33/34 in both cohorts (e.g., in 1958, age 23 vs 33: 9.6% vs 7.8%). Hence, the improvement in the mean of psychological distress between these ages (see Figure 1) was mainly due to a greater decline in the proportion of those with 1-3 symptoms (e.g., in 1958, 23 vs 33: 46.4% vs 38.2%), with a simultaneous increase in the proportion of those with 0 symptoms (e.g., in 1958, 23 vs 33: 44.0% vs 54.0%) (see Figure 2).

**Figure 2.**
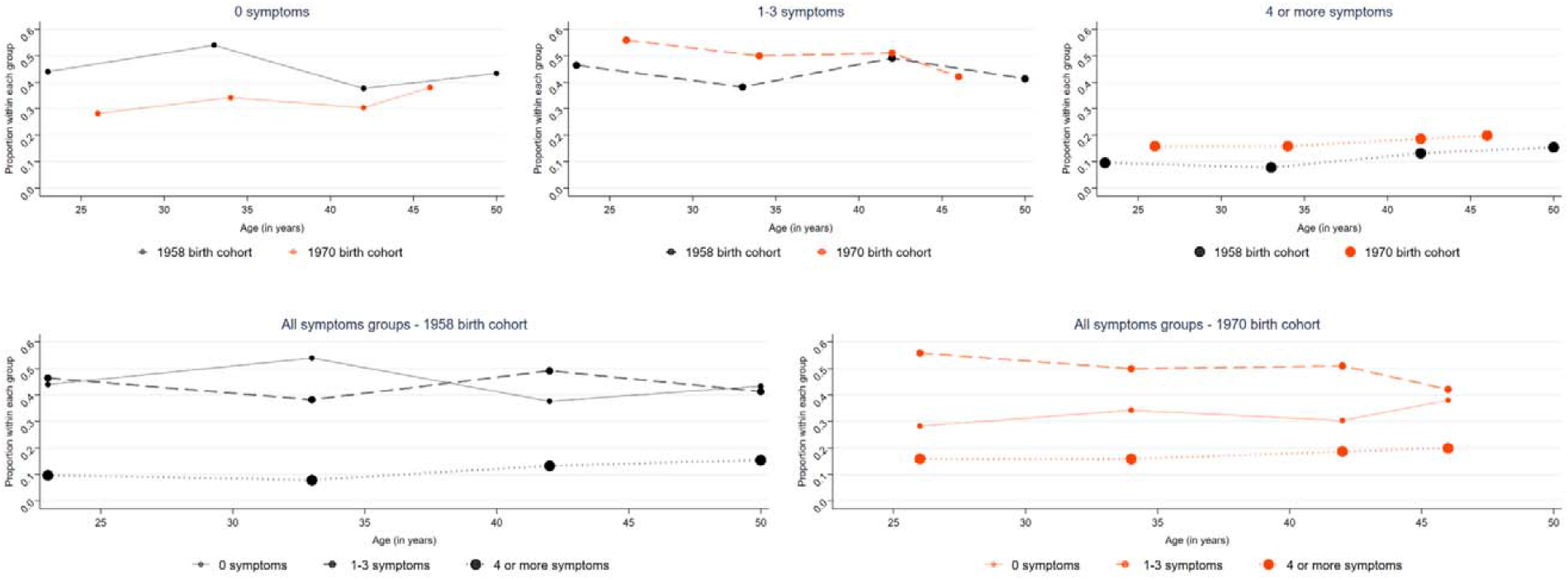
Cohort-stratified age distribution of participants with a varying number of symptoms. *Note*. The figure represents proportion of individuals with varying number of symptoms across age groups and cohorts. Cohort-stratified graphs with all groups of symptoms are given on the same axis to facilitate interpretation of age effects. This graph is an adapted form of a graph in Gondek et al 2021.

Subsequently, it appears that the increase in psychological distress between age 33/34 and 42 was driven by both an increase in the proportion of those with 1-3 symptoms (e.g., in 1958, 33 vs 42: 38.2% vs 49.1%) and 4+ symptoms (e.g., in 1958, 33 vs 42: 7.8% vs 13.2%). The proportion of those with 0 symptoms declined (e.g., in 1958, 33 vs 42: 54.0% vs 37.7%) (see Figure 2).

After age 42, we observed both a higher proportion of those with 0 symptoms (e.g., in 1958, 42 vs 50: 37.7% vs 43.3%) and 4+ symptoms (e.g., in 1958, 42 vs 50: 13.2% vs 15.4%), and a lower proportion of those with 1-3 symptoms (e.g., in 1958, 42 vs 50: 49.1% vs 41.3%) – resulting in polarisation in distress (see Figure 2).

#### Cohort differences (1958 vs 1970) at each age

Members of the 1970 cohort experienced higher psychological distress than those of 1958 across all ages, with the greatest difference at age 33/34 (mean difference: 0.69, 95%CI 0.64 to 0.73). The 1970 cohort also had a greater spread of distribution across all ages, with the difference being the greatest at age 33/34 (standard deviation, 1970 vs 1958: 1.91 vs 1.55) (see Figure 1).

There was a lower proportion – across all ages – of individuals who did not experience any symptoms in the 1970 cohort compared with 1958, with a largely stable proportion of those with one symptom, and an increase across the distribution of those with at least two symptoms (see Figure 2). For instance, at age 33/34 34.2% experienced no symptoms in 1970 compared with 54.0% in the 1958 cohort, whereas 42.3% endorsed at least two symptoms in the 1970 cohort vs 24.7% in 1958. Hence, the entire distribution of psychological distress shifted towards the more severe end of the spectrum in the 1970 cohort compared with the 1958.

### Longitudinal distribution of psychological distress

Psychological distress was higher in the 1970 cohort than in 1958 throughout all ages (23/26 – 46-48/50) (see eFigure 3) – with the mean difference between cohorts reducing with age (age 23/26 vs 46-48/50: 0.55, 95%CI 0.51 to 0.60 vs 0.30, 95%CI 0.25 to 0.36). There was a large spread in the span of trajectories of psychological distress (see eFigure 4). This was mainly due to large between-individuals variance in intercept (centered at age 23/26), which was greater in 1970 than in 1958 cohort (1.74, 95%CI 1.64 to 1.84 vs 1.25, 95%CI 1.19 to 1.31) (see eTable 3 for more details). However, within-individual variance – the fluctuations in distress from occasion to occasion – were greater in the 1970 cohort compared with 1958 (1.52, 95%CI 1.48 to 1.56 vs 1.22, 95%CI 1.19 to 1.24) (see eTable 3). A larger proportion of the total variance in distress was attributed to between-individuals differences in the 1970 cohort than 1958 (53.4% vs 50.6%), whereas a greater amount of variance was explained by within-individual fluctuations in the 1958 cohort (49.4% vs 46.6%).

Due to substantial heterogeneity in the average trajectories of psychological distress, we conducted LCGA in a pooled sample of 1958 and 1970 cohorts identifying four subgroups (classes) of individuals who shared similar trajectories. Comparisons of models with an alternative number of classes pointed towards the model with four classes as the most suitable solution. The four-class solution showed a better fit than solutions with fewer classes (lower values of AIC, BIC and adj BIC; highly significant LMR-LRT and VLMR-LRT) (see eTable 4). In addition, it had a higher entropy than the model with five classes (0.817 vs 0.785) and, as opposed to the solution with five classes, it did not include classes with less than 5% of cohort members. Comparison of cohort-stratified LCGA models also pointed towards the four-class model as the most suitable solution (see eTable 5 for details).

Most participants (72.7%) were in the group with relatively stable “low symptoms”, experiencing on average 0.86 symptoms (standard deviation: 1.16) between age 23/26 – 46-48/50 (see Figure 4). Two other classes with largely stable symptomatology were identified, “moderate symptoms” (with 12.4% of participants; mean: 2.94, standard deviation: 1.69) with stable symptoms until age 42 and a subsequent marginal decrease in symptoms, and “high symptoms” (with 5.7% of participants; mean: 5.14, standard deviation: 2.22), with stable symptoms until age 33/34 and a subsequent marginal increase in symptoms. The final class, “increasing symptoms in midlife” (with 9.1% of participants), comprised individuals who experienced a sharp increase in the mean number of symptoms between age 33/34 and 46-48/50.

The risk of being in any of the symptomatic classes (compared with “low symptoms” group) was higher in the 1970 birth than 1958, by 86% (95%CI 1.74 to 1.99) in relative terms for “moderate symptoms”, 2.09 times% (95%CI 1.88 to 2.31) for “high symptoms”, and 10% (95%CI 1.02 to 1.19) for “increasing symptoms in midlife” (see eTable 6).

**Figure 3.**
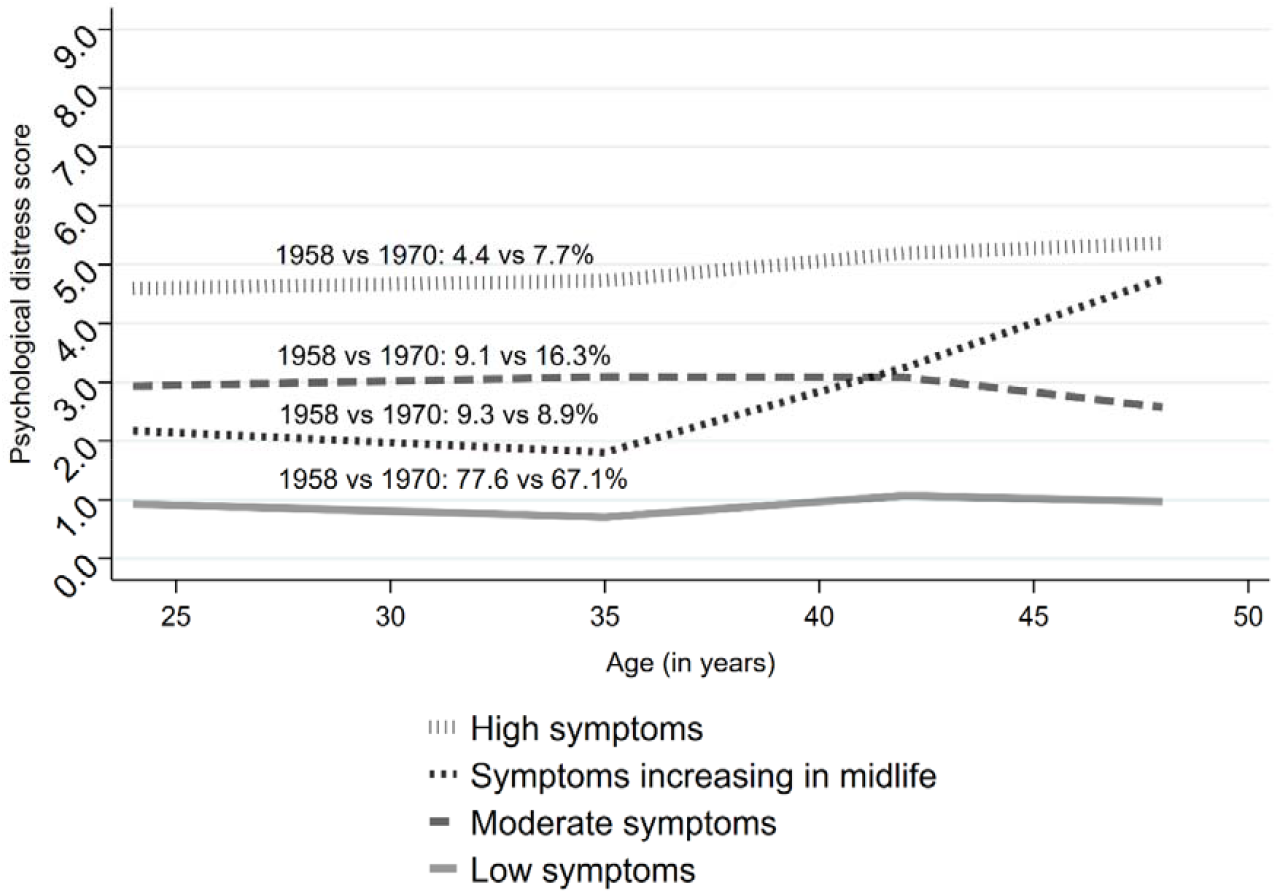
Mean psychological distress across the subgroups of age trajectories of psychological distress, identified with the 4-classes LCGA. *Note*. The figure represents mean psychological distress across age for each subgroup of age trajectories as identified by the final-solution LCGA model. In addition, the prevalence of each class across cohorts is given.

### Demographic composition of subgroups of distress trajectories

The “low symptoms” subgroup was more likely to comprise men (vs women) by 20% (95%CI 1.18 to 1.22) in relative terms, individuals with a high social class at birth (vs low) by 11% (95%CI 1.08 to 1.13), and cohort members with high (vs low) attained qualification by age 30/33 by 21% (95%CI 1.18 to 1.24). There was no evidence for cross-cohort differences in the demographic composition of the “low symptoms” subgroup (see Figure 5 & eTable 6).

The “moderate symptoms” and “high symptoms” subgroups were more likely to include women than men and individuals with low (vs high) social class at birth and low (vs high) qualification attained by age 30/33 (see Figure 4). However, the composition of these subgroups shifted in the 1970 cohort compared with 1958, with a greater proportion of men and those with medium and high qualifications attained by age 30/33 in symptomatic groups. For instance, women were 2.77 times (risk ratio: 2.77, 95%CI 2.31 to 3.32) more likely to be in the “high symptoms” subgroup in the 1958 cohort compared with 1.63 times (95%CI 1.63 to 1.86) in 1970 cohort. For those with low (vs high) qualification attained by age 30/33, the risk of being in the “high symptoms” subgroup was 4.94 times (95%CI 3.63 to 6.72) higher in the 1958 cohort compared with 2.43 (95%CI 2.00 to 2.96) in 1970 cohort. We found no cross-cohort differences due to social class at birth.

**Figure 4.**
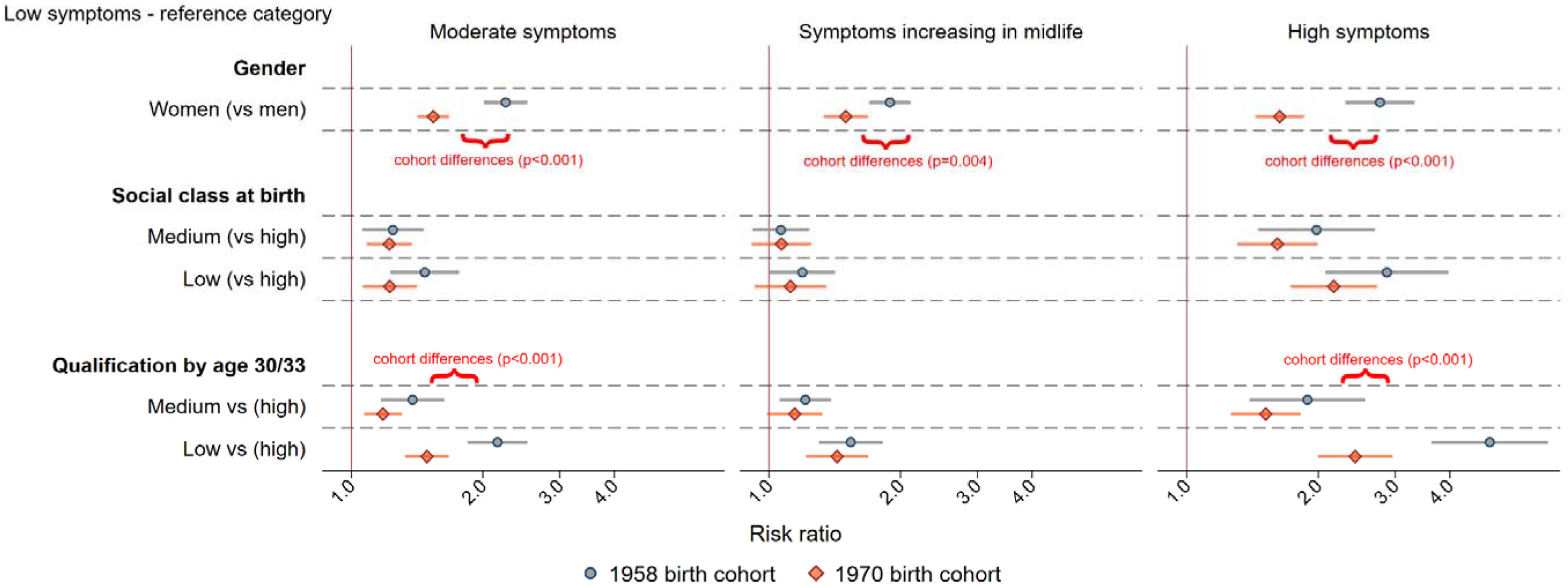
Demographic characteristics of different subgroups of age trajectories of psychological distress – across 1958 and 1970 cohorts. *Note*. The figure represents estimates from Poisson regression using “low symptoms” as a reference category. Curly brackets represent strong evidence for cross-cohort differences in the composition of a given class, indicated as p<0.001 according to a Wald test of a cohort*exposure interaction term. Risk ratio represents relative difference in proportions.

The “symptoms increasing in midlife” subgroup was more likely, to a similar extent in both cohorts, to include individuals with low attained qualification by age 30/33 compared with high (e.g., in 1958 cohort: risk ratio: 1.54, 95%CI 1.30 to 1.82). The likelihood of women being in this subgroup, compared with the “low symptoms”, was lower in the 1970 cohort (risk ratio: 1958 vs 1970: 1.89, 95%CI 1.69 to 2.11 vs 1.52, 95%CI 1.26 to 1.82).

### Symptom level analysis

The prevalence of most symptoms tended to increase in 20s and 40s across both cohorts. The most prevalent symptoms at both ages were worry (age 23/26 vs 33/34 vs 42 – 1958 and 1970 combined: 48.4% vs 39.1% vs 50.2%), feeling upset or irritated (26.4% vs 19.4% vs 25.5%), and low mood (18.8% vs 14.4% vs 20.9%). Feeling of rage was most prevalent in 20s and reduced with age (age 23/26 vs 33/34 vs 42: 7.1% vs 5.2% vs 4.3%), whereas several other symptoms were most prevalent in peoples’ 40s, including fatigue (25.5% vs 25.9% vs 36.7%), nervousness (3.6% vs 3.4% vs 7.7%), and tension (4.4% vs 4.4% vs 6.9%).

We observed a higher prevalence of all symptoms in the 1970 cohort compared with 1958 (see eFigure 5) – particularly worry (mean across all ages: 50.7% vs 41.0%), fatigue (36.8% vs 24.2%), feeling upset or irritated (28.9% vs 21.4%), low mood (21.0% vs 16.4%), and nervousness (4.5% vs 8.1%). There were more modest increases in the prevalence of panic (7.5% vs 9.8%), feeling scared (7.3% vs 9.0%), tension (5.4% vs 6.8%), and rage (4.7% vs 5.5%).

## Discussion

### Summary of main findings

The spread of distribution in distress increased with age, with the shifts in the distribution mainly due to changing proportion of those with moderate symptoms, except for midlife (age 42-50) when we observed polarisation in distress – increased proportions of people with and without multiple symptoms. We found large variance around population-average age trajectories of distress, identifying four subgroups of similar age profiles of distress. These were characterised by low symptoms, moderate symptoms, symptoms drastically increasing in midlife, and high symptoms.

The elevated levels of distress in the 1970 cohort, compared with the 1958 cohort, appeared to be due to an increase in the proportion of individuals with both moderate and high symptoms. These observed cohort differences were driven to some extent by an increasing proportion of men and individuals with high qualification in the moderate and high distress groups.

### Interpretation and implications of the findings

Considering distributional shifts in the population across age or cohorts is important for public health policy. It helps us understand who drives the trends over time, which in turn, may facilitate monitoring of these populations at risk, understanding risk factors that they are affected by and putting them into a greater focus of policies or interventions. Based on the observed shifts in the distribution of distress, we speculate on circumstances contributing to these shifts across ages and cohorts.

### Age differences across cohorts (1958 and 1970)

In line with previous evidence [2], we found a large spread in distribution of distress at any given age. Despite a distinct population-average age trend in distress, as revealed by the multilevel regression analysis, age trajectories tend to be highly variable and individualised. Hence, it has been advocated that we aim to uncover diversity in distributions of distress or wellbeing, which could facilitate understanding what leads to deviations from average [32]. In the current study, we explored this heterogeneity both cross-sectionally and longitudinally.

First, we compared shifts in the cross-sectional distribution of distress among those with a varying number of symptoms. A particularly striking finding was that whilst higher distress in mid-20s and early-40s was driven by shifts in the entire distribution, after age 42 we observed a polarisation in distress – with a simultaneous increase in the proportion of those with no symptoms and high symptoms. As the mean of distress remained largely stable between age 42 and 46/50, this trend would be masked when examining mean differences. We speculate that as people approach midlife, they may share certain stressors that affect the population across the distribution. However, as they progress through midlife some might be better equipped to deal with these stressors, whereas others experience further increases in distress. This diverging trend can be explained by midlife being described both as the life phase of considerable challenges and opportunities [33, 34]. Midlife tends to be characterised by an increase in intensity and load of multiple concurrent stressors, including caring for ageing parents and children, managing peaking careers, and dealing with early signs of declining health [33, 34]. These factors can help to explain a particularly high prevalence of symptoms such as fatigue, nervousness, and tension during this life phase [5]. Concurrently, midlife also tends to be a peak time for earnings, self-confidence, and responsibility across various areas of life [33, 34], with some being better equipped to deal with midlife adversity, utilising available resources to a greater extent.

Secondly, we aimed to statistically identify subgroups of individuals sharing comparable longitudinal trajectories. Majority of individuals were classed as having low psychological distress throughout adulthood, which can be considered as the typical developmental trajectory. Two other groups of particular interest were characterised by persistently high distress, which marginally increased from their mid-30s, and initially moderate levels of symptoms with a sharp increase from mid-30s. Importantly, these subgroups of trajectories were consistently identified across both birth cohorts, despite experiencing the same age at different historical time. This would suggest that age effects play a more important role in explaining these trajectories than historical context. For instance, members of both cohorts had particularly high distress at age 42, in the year 2000 for those born in 1958 and year 2012 for those born in 1970, despite these two periods being largely different. For instance, the economy was soaring in 2000, with widespread employment opportunities (e.g., unemployment was around 5%), whereas 2012 was in stark contrast, with the economy going through stagnation and unemployment reaching around 8% [35]. Future research could examine what makes certain groups particularly susceptible for midlife distress, compared to those with low symptoms, while focusing on experiences that are typically associated with a given life phase regardless a birth cohort being born into. For instance, this could include work factors (e.g., increasing job responsibilities), family factors (e.g., increasing care demands), declining physical health or adverse life events (e.g., divorce) [36].

Cohort differences (1958 vs 1970) across age

The entire distribution of distress shifted towards a more severe end of the spectrum in 1970, compared with the 1958 cohort. We did not observe polarisation in distress over time previously found in the younger population (16-24 years old), between 1991 and 2008, in the British Household Panel Survey [11]. ‘Generation X’ (born between 1961-1981), compared with ‘Baby boomers’ (1946-1964), experienced a plethora of risk factors strongly associated with mental health, such as rising inequality, unemployment among young people in the mid-1980s, or changes in family structure (e.g., increasing rates of divorce) [37], which may explain why the more recent cohort experienced an increase in distress across the entire distribution of distress and all studied symptoms.

Interestingly, it appears that members of the 1970 birth cohort experience not only higher levels but also greater instability in distress, as implied by within-person variance. There is a plethora of potential reasons for this phenomenon. This can be due to a range of social, behavioural and environmental factors leading to fluctuations of distress within the individual. For instance, changes in employment (e.g., losing or getting a job), or family situation (e.g., getting married, moving a house or getting divorced) may result in within-person instabilities in distress. These experiences tend to be more common among individuals born around 1970, hence possibly explaining greater variance in distress in this cohort [37].

Gender and socioeconomic inequalities in psychological distress reduced in magnitude due to a greater increase in the proportion of men and those with a medium and high attained qualification in symptomatic groups. The findings that the mental health of women did not worsen as much as of men in the 1970 birth cohort compared with 1958, may be due to achieving equal levels of school success (e.g., in obtaining a university degree) and improved labour market opportunities among women [37]. Moreover, economic downturns, which have been found to affect men’s mental health to a greater extent [38], as well as dispersion of traditional manufacturing mainly employing men [37], may partially explain a greater increase in distress among men in this more recent cohort. Reporting mental health problems might have also become more acceptable among men, who have also become increasingly involved in household chores and balancing work and childcare, which might have more negatively affected their mental health in comparison to women [39]. However, more recent cohort trends, particularly among young adults, suggest that the gender gap may be widening again [4, 11].

Socioeconomically advantaged individuals experienced a greater increase in distress in the more recent cohort. This is a somewhat surprising finding in the context of rising economic inequality over time, which one would expect to benefit more advantaged groups [37]. However, social inequality has been linked with poor social cohesion, which is associated with higher levels of distress among those from both advantaged and disadvantaged socioeconomic groups [40]. In addition, some have posited that rising house prices have affected relatively well-off members of the middle class, who tend to work increasingly longer hours, borrow more, commute longer and save less to keep up with this increase – all of which may contribute to worsening mental health among those with high qualification in more recent cohorts [41].

Increases in distress in the more recent birth cohort, which disproportionally affected more socioeconomically advantaged groups, warrant policies that focus is on improving population mental health on the entire socioeconomic spectrum, while aiming to also reduce socio-economic inequalities. It has been widely advocated that policy efforts should focus on the most vulnerable groups to bridge the socioeconomic gap in mental health, for instance, by protecting the employment rights of those on insecure contracts, or poorly paid part-time workers [12]. However, it is equally important to ensure positive psychosocial conditions, aiming to reduce unhealthy stress, in jobs typically considered of higher social classes [12].

### Strengths and limitations

The main strength of our study is that it is broadly representative of British men and women born around 1958 and 1970. However, both cohorts suffered from missing data due to non-response and attrition, which may have introduced bias. This limitation was mitigated by FIML and multiple imputation, while taking advantage of the rich information available in both cohorts to minimise bias in estimates [42]. The study is limited by the gaps between assessments in the two cohorts, more frequent assessments would have permitted a more age nuanced picture of the changing distributions across cohorts. Similarly, comparing a greater number of generations would also provide a fuller picture of long-term trends, however, long-term longitudinal studies across multiple generations are limited in availability. As recent cohorts highlight further increases in distress at younger ages, examining these distributions as they move into adulthood will be valuable [43].

Another limitation of our study is that psychological distress was self-reported, rather than being ascertained by a clinical interview. However, the Malaise Inventory was deemed to be an appropriate measure for the objectives of the study, which were not to provide descriptive epidemiology of psychological distress but rather compare its distribution across various sociodemographic groups. The key limitation of the Malaise Inventory in the context of our study is that it aims to describe current levels of psychological distress, hence it may be prone to influences by recent life circumstances resulting in large within-individual variance. This, however, should not bias cross-cohort comparisons. Further, although short measures of symptoms are widespread in population-based research, they do not include a comprehensive range of possible symptomology in the population and studies using more detailed measures might provide greater insights into the specific symptoms that are salient in changing lifecourse and cohort trends.

## Conclusion

Midlife appears to have a polarising effect on the experience of distress, with a rising proportion of individuals with both no symptoms and high symptoms. We also found that the 1970 cohort, compared to 1958, experienced a shift in the entire distribution of distress towards the more severe end of the spectrum. Our study demonstrates the importance of studying not only mean levels of distress over time, but also the underlying shifts in its distribution. Due to the large dispersion of distress scores at any given measurement occasion, understanding the underlying distribution provides a more complete picture of the population trends.

## Data Availability

Cohort data comply with ESRC data sharing policies, readers can access data via the UK Data Archive (www.data-archive.ac.uk), through a formal request.

## Acknowledgements

We thank the Centre for Longitudinal Studies (CLS), UCL Institute of Education, for the use of these data and the UK Data Service for making them available. Neither CLS nor the UK Data Service bear any responsibility for the analysis or interpretation of these data.

## Author contributions

All authors contributed to the study conception and design. Analysis was performed by Dawid Gondek. The first draft of the manuscript was written by Dawid Gondek and all authors commented on previous versions of the manuscript. All authors read and approved the final manuscript.

## Ethical Standards

The authors assert that all procedures contributing to this work comply with the ethical standards of the relevant national and institutional committees on human experimentation and with the Helsinki Declaration of 1975, as revised in 200.

## Patient consent

All participants provided written informed consent after a thorough explanation of the research procedures.

The lead author (Dawid Gondek) affirms that this manuscript is an honest, accurate, and transparent account of the study being reported; that no important aspects of the study have been omitted; and that any discrepancies from the study as planned have been explained.

## Dissemination to participants and related patient and public communities

The results of this research will be reported in newsletters for study participants, and public lectures.

## Patient involvement

This research was done without patient involvement. Patients were not invited to comment on the study design and were not consulted to develop patient relevant outcomes or interpret the results. Patients were not invited to contribute to the writing or editing of this document for readability or accuracy.

