## Supplemental Material for "How is the distribution of psychological distress changing over time? Who is driving these changes? Analysis of the 1958 and 1970 British birth cohorts"

### eFigure 1. Flow diagram of the study sample.

**Original sample**

**(born in Great Britain)**

1958 birth cohort: n = 17,415

1970 birth cohort: n = 17,196

**Died or emigrated by the last data sweep** 1958 birth cohort (age 50): n = 2,578

1970 birth cohort (age 46-48): n = 1,375

**Did not have any measure of distress**

1958 birth cohort (age 50): n = 1,587

1970 birth cohort (age 46-48): n = 4,364

1958 birth cohort: n = 14,837

1970 birth cohort: n = 15,821

**Analytical sample**

1958 birth cohort: n = 13,250

1970 birth cohort: n = 11,457

Total: n = 24,707

### eTable 1. Items of the Malaise Inventory.

How are you feeling generally…

1. Do you often have backache?

**2. Do you feel tired most of the time?**

**3. Do you often feel miserable or depressed?**

4. Do you often have bad headaches?

**5. Do you often get worried about things?**

6. Do you usually have great difficulty in falling or staying asleep?

7. Do you usually wake unnecessarily early in the morning?

8. Do you wear yourself out worrying about your health?

**9. Do you often get in a violent rage?**

10. Do people often annoy and irritate you?

11. Have you at times had twitching of the face, head or shoulders?

**12. Do you often suddenly become scared for no good reason?**

13. Are you scared to be alone when there are no friends near you?

**14. Are you easily upset or irritated?**

15. Are you frightened of going out alone or of meeting people?

**16. Are you constantly keyed up and jittery?**

17. Do you suffer from indigestion?

18. Do you suffer from an upset stomach?

19. Is your appetite poor?

**20. Does every little thing get on your nerves and wear you out?**

**21. Does your heart often race like mad?**

22. Do you often have bad pains in your eyes?

23. Are you troubled with rheumatism or fibrositis?

24. Have you ever had a nervous breakdown?

*Note*. In bold – nine items used across all ages (23-50).

### eAppendix 1 Missing data strategy.

Most participants had at least two measures of psychological distress – n=11,558 (87.2% of the total included sample) in the 1958 birth cohort and n=9,224 (80.51% of the total included sample) in the 1970 birth cohort. In both birth cohorts, similar characteristics were predictive of having missing information, these were being a man, being a more disadvantaged social class at birth, having lower birthweight, having higher psychological distress, mother not being married at birth, a mother being a teen at birth, mother ever smoking during pregnancy (see eTable 2).

For the cross-sectional analyses, the missing data were imputed using multiple imputation by chained equations (MICE), due to the non-monotone pattern of missing values, and due to its ability to accommodate various types of variables in the imputation model, including continuous and categorical ones. This approach uses a series of univariate conditional imputation models to impute missing data.^1^

The imputation model included all individual items of the Malaise Inventory, indicators of gender, father’s occupational class at birth, birthweight (in kg), mother being married at birth, mother’s age at birth (in years), mother ever smoking during birth, highest achieved professional qualification by age 30/33. For the analyses with total scores on the Malaise Inventory, the individual items were summed up after the imputation. It is recommended to impute variables in the form, in which they are going to be used in the analysis to preserve the relationship between the variables.^2,3^ Hence, we also conducted multiple imputation with total scores of the Malaise Inventory. Both approaches resulted in highly comparable results, hence for simplicity, we used the datasets based on the imputation of individual items for all cross-sectional analyses.

The multiple imputation works under the assumption that the data are missing at random (MAR).^4,5^ The MAR mechanism, which is largely untestable, implies that systematic differences between the missing and the observed values can be explained by observed data.^4^ The plausibility of the MAR assumption was maximised by including auxiliary variables, improving the accuracy of the MI and minimising non-random variation in the imputed values.^6,7^ These variables included gender, father’s occupational class at birth, birthweight (in kg), indicators of mother being married at birth, mother’s age at birth (in years), mother ever smoking during birth.

As shown by eTable 2, these variables were strongly associated with having missing data and with psychological distress at age 46-48/50. In addition, the imputation model was rich as psychological distress (i.e., individual items of the Malaise Inventory) at the preceding collection wave was a strong predictor of having missing data at the subsequent collection waves.

The longitudinal analyses, including multilevel linear regression and latent class growth analysis were conducted using Full Information Maximum Likelihood (FIML). FIML accounts for missing information, hence individuals with at least one measure of psychological distress were included. Likewise, multiple imputation FIML works under the assumption that data are MAR. Hence, LCGA was enriched by adding auxiliary variables that predict missingness and the outcome (gender, father’s occupational class at birth, birthweight (in kg), indicators of a mother being married at birth, mother’s age at birth (in years), mother ever smoking during birth).^8^

| eTable 2. Predictors of having missing data on psychological distress at any point (vs not having missing data) and of psychological distress at age 50. | | | | |
| --- | --- | --- | --- | --- |
|  | 1958 birth cohort | | 1970 birth cohort | |
|  | Missing | Distress (age 50) | Missing | Distress (age 46-48) |
|  | RR (95% CI) | B (95% CI) | RR (95% CI) | B (95% CI) |
| Father’s occupational class at birth  (I – professional – reference) |  | |  |  |
| II – Managerial and technical | 0.91 (0.82 to 1.01) | 0.04 (-0.17 to 0.26) | 1.06 (0.98 to 1.16) | 0.14 (-0.10 to 0.36) |
| III – Skilled nonmanual/manual | 1.10 (1.00 to 1.21) | 0.22 (0.03 to 0.41) | 1.17 (1.08 to 1.26) | 0.25 (0.04 to 0.45) |
| IV – Partly-skilled | 1.14 (1.03 to 1.27) | 0.33 (0.11 to 0.54) | 1.31 (1.22 to 1.42) | 0.36 (0.12 to 0.61) |
| V – Unskilled | 1.34 (1.22 to 1.49) | 0.45 (0.22 to 0.68) | 1.40 (1.29 to 1.52) | 0.41 (0.11 to 0.72) |
| Men (women – reference category) | 1.13 (1.09 to 1.17) | -0.59 (-0.67 to -0.51) | 1.13 (1.10 to 1.16) | -0.48 (-0.58 to -0.39) |
| Birthweight (in kg) | 0.96 (0.93 to 0.99) | -0.16 (-0.23 to -0.08) | 0.95 (0.93 to 0.98) | -0.20 (-0.29 to -0.10) |
| Mother married at birth (no – reference category) | 1.21 (1.12 to 1.31) | 0.25 (0.03 to 0.48) | 1.15 (1.10 to 1.20) | 0.48 (0.26 to 0.70) |
| Mother’s age at birth (<19 – reference category) | 1.16 (1.09 to 1.24) | 0.20 (0.02 to 0.38) | 1.15 (1.11 to 1.19) | 0.26 (0.08 to 0.44) |
| Mother smoked during birth  (no – reference category) | 1.12 (1.08 to 1.16) | 0.16 (0.08 to 0.25) | 1.10 (1.07 to 1.13) | 0.24 (0.14 to 0.34) |
| Psychological distress at age 42 | 1.04 (1.03 to 1.05) | 0.64 (0.62 to 0.66) | 1.03 (1.02 to 1.05) | 0.69 (0.67 to 0.71) |
| *Note.* RR = risk ratio; B = beta coefficient. | | | | |

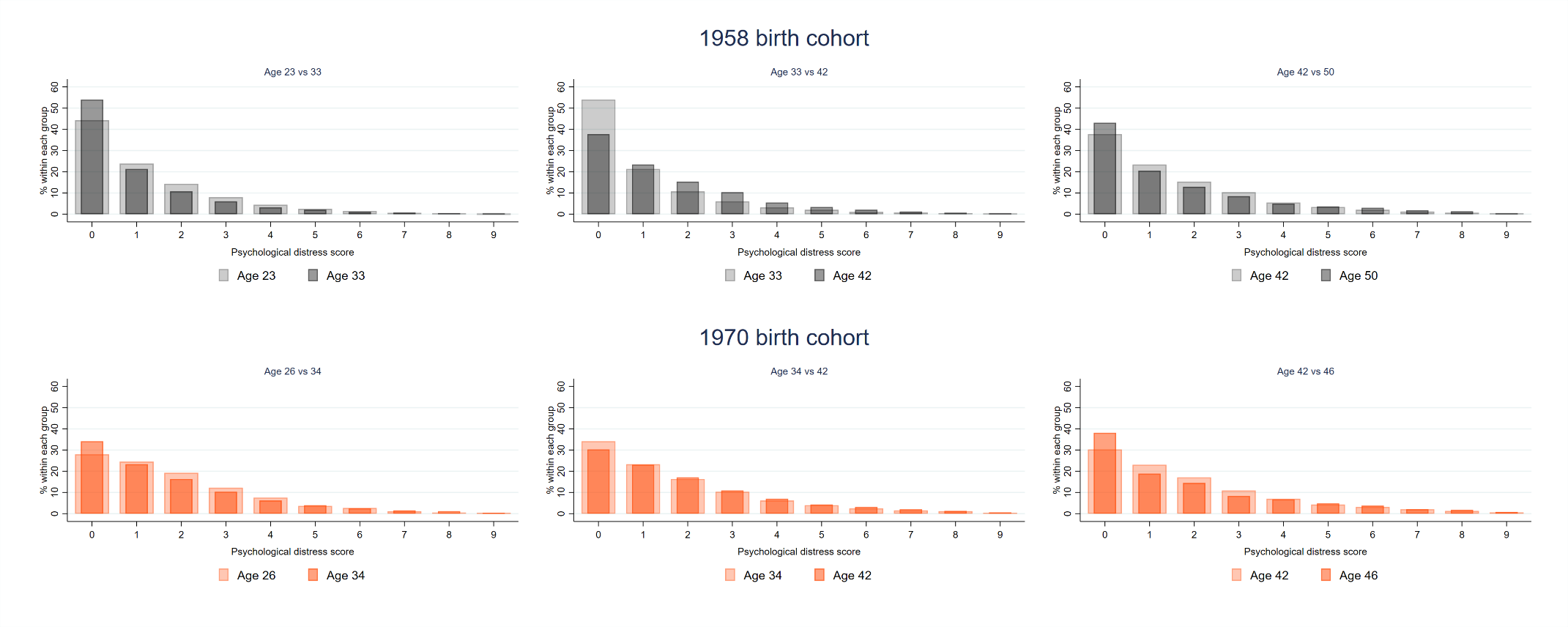

### eFigure 2. Cross-sectional distribution of psychological distress across age within each cohort.

### eAppendix 2 Trajectories of distress – modelling strategy.

We built the multilevel regression models starting with including age polynomials up to a cubic term and retaining them if their coefficients did not equal zero, according to the Wald test (at p<0.05), and their inclusion improved the model fit according to the likelihood ratio test (at p<0.05). This was due to the previously found non-linearity of the growth of psychological distress in adulthood.^9^ The fixed part of all models included age terms and the intercept (age 23/26). The random part of the model captured variance in the intercept and age slope, allowing for heterogeneity in the trajectories of psychological distress. The model was stratified by cohorts (1958 and 1970).

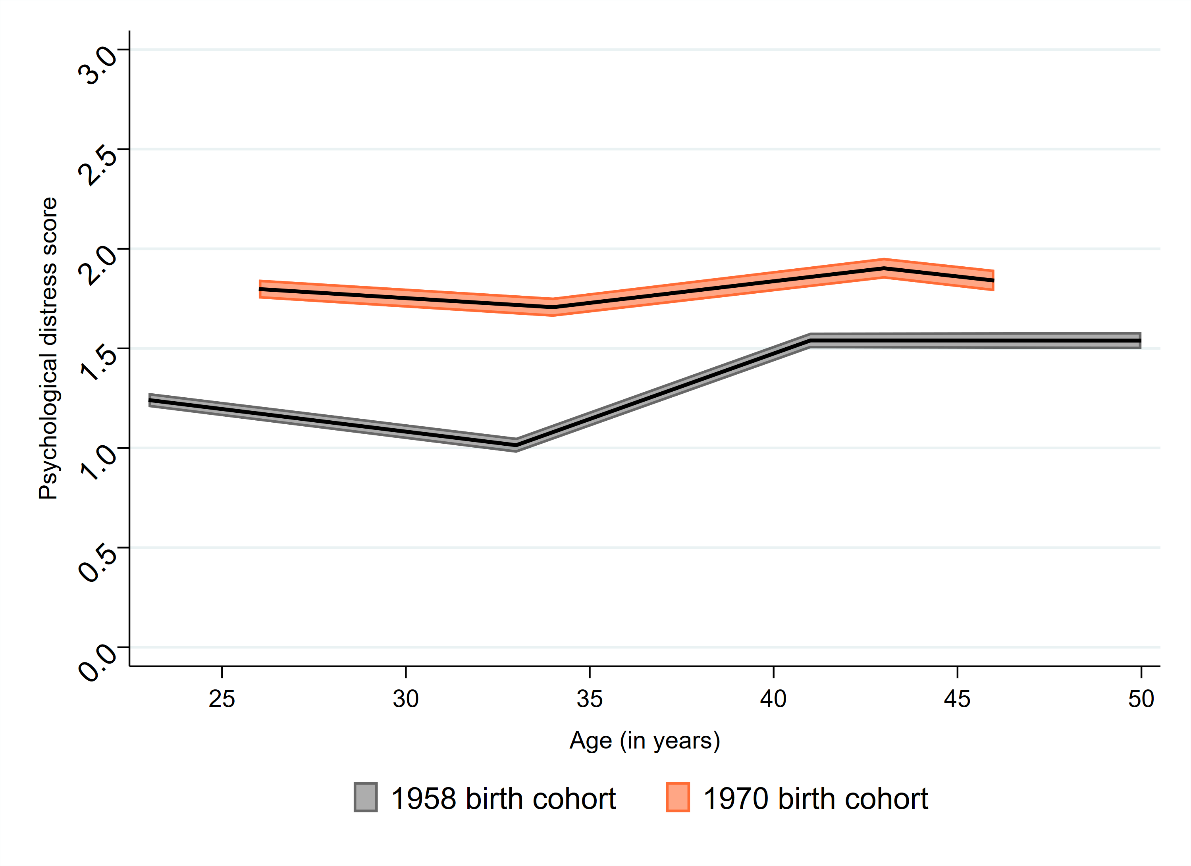

### eFigure 3. Age trajectory of psychological distress – estimates from multilevel linear regression.

*Note.* The trajectories are based on the estimates from eTable 3.

| eTable 3. Age trajectory of psychological distress – estimates from multilevel linear regression. | | | | |
| --- | --- | --- | --- | --- |
|  |  | **1958 birth cohort** |  | **1970 birth cohort** |
| *Fixed effects* |  | β (95% CI) |  | β (95% CI) |
| Intercept (centered at age 23/26) |  | 1.24 (1.22 to 1.27) |  | 1.81 (1.77 to 1.85) |
| Age |  | -0.12 (-0.13 to -0.11) |  | -0.06 (-0.08 to -0.05) |
| Age^2^ |  | 0.01 (0.01 to 0.01) |  | 0.01 (0.01 to 0.01) |
| Age^3^ |  | -0.00 (-0.00 to -0.00) |  | -0.00 (-0.00 to -0.00) |
| *Random effects* |  | σ² (95% CI) |  | σ² (95% CI) |
| Intercept variance |  | 1.25 (1.19 to 1.31) |  | 1.74 (1.64 to 1.84) |
| Slope variance |  | 0.00 (0.00 to 0.00) |  | 0.00 (0.00 to 0.00) |
| Intercept-slope covariance |  | 0.00 (-0.00 to 0.00) |  | 0.01 (0.01 to 0.01) |
| Within-individual variance |  | 1.22 (1.19 to 1.24) |  | 1.52 (1.48 to 1.56) |
| *Variance Partitioning Coefficient* |  |  |  |  |
| Age (level 2) |  | <0.01 |  | <0.01 |
| Individual (level 2) |  | 50.6 |  | 53.4 |
| Observation (level 1) |  | 49.4 |  | 46.6 |
| Observations |  | 41,177 |  | 31,446 |
| Participants |  | 13,250 |  | 11,457 |
| *Note*. Variance Partitioning Coefficient (VPC) represents the proportion of variation attributed to each random level. | | | | |

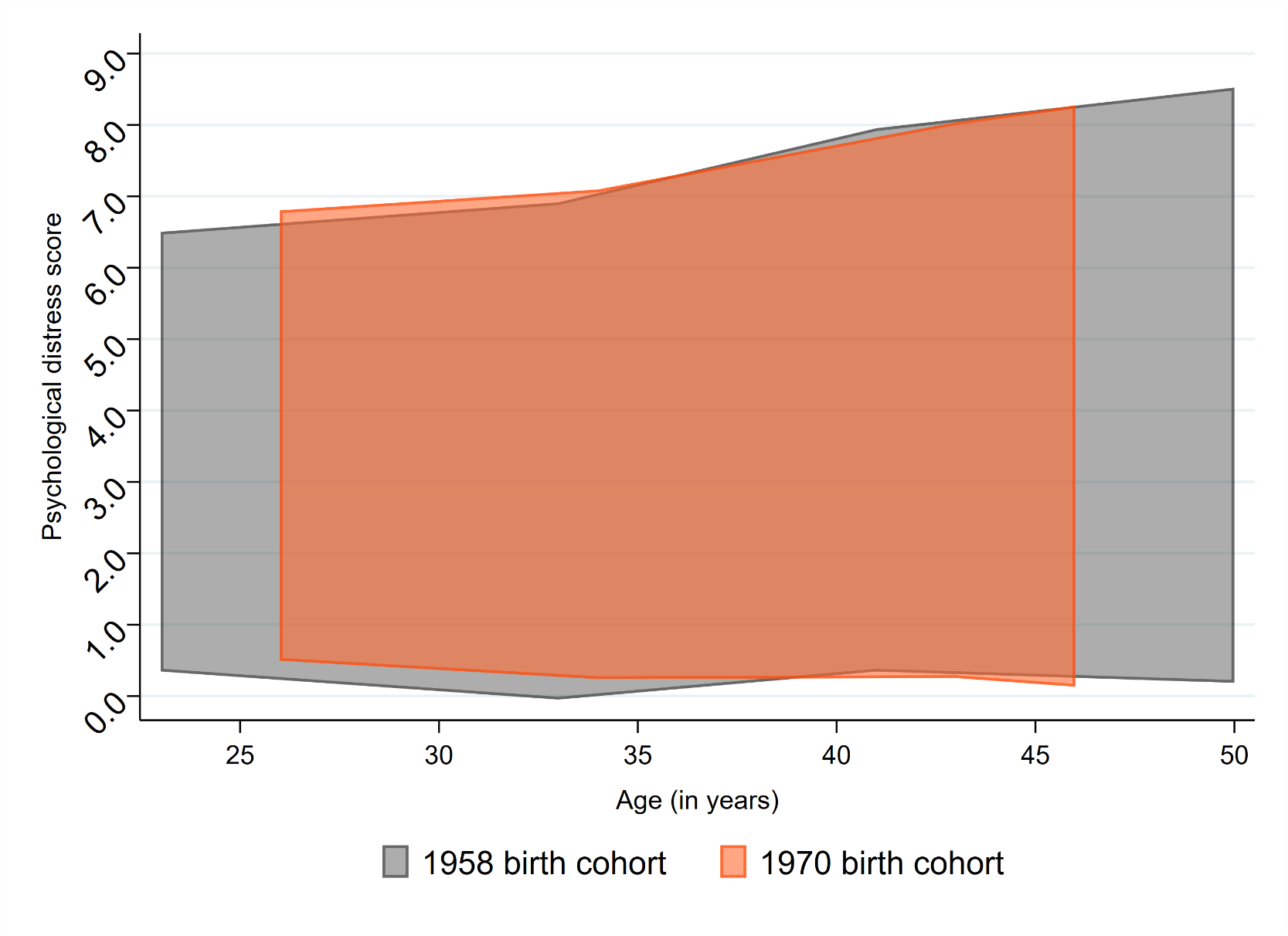

### eFigure 4. Span of individual age trajectories of psychological distress.

*Note.* These show the extents to which individuals vary in their trajectories of psychological distress in 1958 and 1970 cohorts. The trajectories are based on the estimates from eTable 3.

| eTable 4. Comparison of models with different numbers of classes obtained with latent class growth analysis. | | | | |
| --- | --- | --- | --- | --- |
|  | **2 classes** | **3 classes** | **4 classes** | **5 classes** |
| AIC | 270140.183 | 264664.635 | 261070.739 | 259268.122 |
| BIC | 270229.446 | 264786.358 | 261224.921 | 259454.764 |
| Sample-adjusted BIC | 270194.488 | 264738.688 | 261164.539 | 259381.670 |
| Entropy | 0.868 | 0.826 | 0.817 | 0.785 |
| LMR-LRT | <0.00001 | <0.00001 | <0.00001 | <0.00001 |
| VLMR-LRT | <0.00001 | <0.00001 | <0.00001 | <0.00001 |
| % of the smallest class | 17.7% | 6.5% | 5.7% | 4.5% |
| *Note*. AIC = Akaike Information Criterion; BIC = Bayesian Information Criterion; (V)LMR-LRT = (Vuong) Lo–Mendell–Rubin likelihood ratio tests. | | | | |

| eTable 5. Comparison of models with different numbers of classes obtained with latent class growth analysis. | | | | | | | | |
| --- | --- | --- | --- | --- | --- | --- | --- | --- |
|  | **1958** | **1970** | **1958** | **1970** | **1958** | **1970** | **1958** | **1970** |
|  | **2 classes** | | **3 classes** | | **4 classes** | | **5 classes** | |
| AIC | 147353.594 | 121105.471 | 143931.603 | 118842.941 | **141734.302** | **117622.299** | 140396.645 | 116915.654 |
| BIC | 147436.003 | 121186.281 | 144043.980 | 118953.137 | **141876.645** | **117761.880** | 140568.955 | 117084.620 |
| Sample-adjusted BIC | 147401.046 | 121151.324 | 143996.311 | 118905.468 | **141816.265** | **117701.500** | 140495.864 | 117011.529 |
| Entropy | 0.895 | 0.838 | 0.850 | 0.794 | **0.842** | **0.780** | 0.841 | 0.750 |
| LMR-LRT | <0.0000 | <0.0000 | <0.0000 | <0.0000 | **<0.0000** | **<0.0000** | 0.0016 | 0.0008 |
| VLMR-LRT | <0.0000 | <0.0000 | <0.0000 | <0.0000 | **<0.0000** | **<0.0000** | 0.0018 | 0.0009 |
| % of the smallest class | 14.9 | 20.3 | 5.6 | 7.7 | **4.9** | **6.4** | 2.8 | 5.4 |
| *Note*. AIC = Akaike Information Criterion; BIC = Bayesian Information Criterion; (V)LMR-LRT = (Vuong) Lo–Mendell–Rubin likelihood ratio tests. | | | | | | | | |

### eFigure 5. Cross-sectional proportions of each symptom across age groups and cohorts (1958 vs 1970).

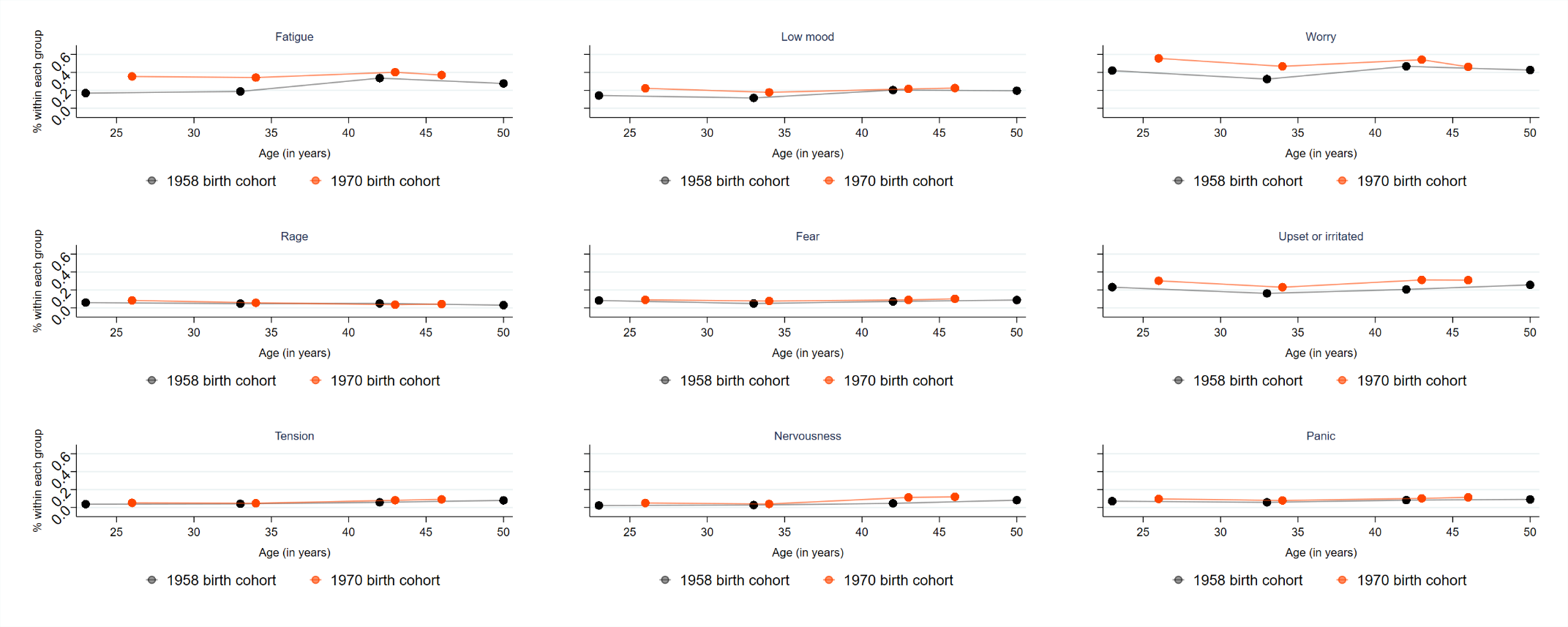

| eTable 6. Demographic composition of the four classes identified by latent class growth analysis – stratified by the 1958 and 1970 birth cohorts. | | |
| --- | --- | --- |
|  | Moderate symptoms (vs low symptoms) | |
|  | 1958 | 1970 |
|  | RR (95% CI) | RR (95% CI) |
| Birth cohort (1958 – reference) |  | 1.86 (1.74 to 1.99) |
| Women (men – reference) | 2.26 (2.01 to 2.53) | 1.54 (1.42 to 1.67) |
| Social class at birth (high I-II – reference) |  |  |
| Medium (III) | 1.25 (1.06 to 1.46) | 1.22 (1.08 to 1.38) |
| Low (IV-V) | 1.47 (1.23 to 1.76) | 1.22 (1.06 to 1.41) |
| Highest qualification at age 30-33  (high NVQ 4-5 – reference) |  |  |
| Medium (NVQ 2-3) | 1.38 (1.17 to 1.63) | 1.18 (1.07 to 1.31) |
| Low (none-NVQ 1) | 2.16 (1.84 to 2.53) | 1.49 (1.33 to 1.67) |
|  | Symptoms increasing in midlife  (vs low symptoms) | |
|  | 1958 | 1970 |
|  | RR (95% CI) | RR (95% CI) |
| Birth cohort (1958 – reference) |  | 1.10 (1.02 to 1.19) |
| Women (men – reference) | 1.89 (1.69 to 2.11) | 1.50 (1.33 to 1.68) |
| Social class at birth (high I-II – reference) |  |  |
| Medium (III) | 1.06 (0.92 to 1.24) | 1.07 (0.91 to 1.25) |
| Low (IV-V) | 1.19 (1.00 to 1.42) | 1.12 (0.93 to 1.35) |
| Highest qualification at age 30-33  (high NVQ 4-5 – reference) |  |  |
| Medium (NVQ 2-3) | 1.21 (1.06 to 1.39) | 1.14 (0.99 to 1.32) |
| Low (none-NVQ 1) | 1.54 (1.30 to 1.82) | 1.43 (1.22 to 1.69) |
|  | High symptoms (vs low symptoms) | |
|  | 1958 | 1970 |
|  | RR (95% CI) | RR (95% CI) |
| Birth cohort (1958 – reference) |  | 2.09 (1.88 to 2.31) |
| Women (men – reference) | 2.77 (2.31 to 3.32) | 1.63 (1.44 to 1.86) |
| Social class at birth (high I-II – reference) |  |  |
| Medium (III) | 1.98 (1.46 to 2.70) | 1.61 (1.30 to 1.99) |
| Low (IV-V) | 2.87 (2.07 to 3.97) | 2.17 (1.73 to 2.73) |
| Highest qualification at age 30-33  (high NVQ 4-5 – reference) |  |  |
| Medium (NVQ 2-3) | 1.89 (1.39 to 2.56) | 1.52 (1.26 to 1.82) |
| Low (none-NVQ 1) | 4.94 (3.63 to 6.72) | 2.43 (2.00 to 2.96) |
| *Note*. RR=risk ratio; CI=confidence interval; NVQ=National Vocational Qualifications. | | |
